# COVID-19 vaccine hesitancy in Australian patients with solid organ cancers

**DOI:** 10.1101/2022.07.08.22277398

**Authors:** N. Bain, M. Nguyen, L. Grech, D. Day, A. McCartney, K. Webber, A. Kwok, S. Harris, H. Chau, B. Chan, L. Nott, N. Hamad, A. Tognela, C. Underhill, B.S. Loe, D. Freeman, E. Segelov, the CANcer patients’ perspectives on coronavirus VACCination Survey(CANVACCS) investigators

**Author notes:** **<u>Corresponding author</u>** Dr Nathan T Bain, Oncology Department, Monash Medical Centre, 246 Clayton Road, Clayton VIC 3168, Australia. NB and MN contributed equally.

## Abstract

**Background:** Vaccination is the cornerstone of the global public health response to the COVID-19 pandemic. Excess morbidity and mortality of COVID-19 infection is seen in people with cancer. COVID-19 vaccine hesitancy has been observed in this medically vulnerable population, although associated attitudes and beliefs remain poorly understood.

**Patients and Methods:** An online cross-sectional survey of people with solid organ cancers was conducted through nine health services across Australia. Demographics, cancer-related characteristics, and vaccine uptake were collected. Perceptions and beliefs regarding COVID-19 vaccination were assessed using the Oxford COVID-19 Vaccine Hesitancy Scale, the Oxford COVID-19 Vaccine Confidence and Complacency Scale, and the Disease Influenced Vaccine Acceptance Scale-6.

**Results:** Between June and October 2021, 2691 people with solid organ cancers completed the survey. Median age was 62.5 years (*SD*=11.8; range 19-95), 40.9% were male, 71.3% lived in metropolitan areas, and 90.3% spoke English as their first language. The commonest cancer diagnoses were breast (36.6%), genitourinary (18.6%) and gastrointestinal (18.3%); 59.2% had localized disease and 56.0% were receiving anti-cancer therapy. Most participants (79.7%) had at least one COVID-19 vaccine dose. Vaccine uptake was higher in people who were older, male, metropolitan, spoke English as a first language, and had a cancer diagnosis for more than six months. Vaccine hesitancy was higher in people who were younger, female, spoke English as a non-dominant language and lived in a regional location, and lower in people with genitourinary cancer. Vaccinated respondents were more concerned about being infected with COVID-19 and less concerned about vaccine safety and efficacy.

**Conclusions:** People with cancer have concerns about acquiring COVID-19, which they balance against vaccine-related concerns about the potential impact on their disease progress and/or treatment. Detailed exploration of concerns in cancer patients provides valuable insights, both for discussions with individual patients and public health messaging for this vulnerable population.

## Introduction

Globally, there have been over 500 million SARS-CoV-2 infections and 6 million deaths during the pandemic as of March 2022 (1). People with cancer are particularly vulnerable due to immunosuppression from the underlying disease and treatments, and comorbid medical conditions (2). A recent systematic review and meta-analysis of 81 studies of 61532 patients reported that people with cancer are at approximately two times greater risk of serious illness and death from COVID-19 compared to the general population (3).

Vaccination is a critical component of the public health response to the pandemic (1). COVID-19 vaccines have demonstrated efficacy in reducing infection, serious illness and death, and evidence is growing for real world effectiveness of community-wide vaccination programs (4–7). Pivotal clinical trials excluded patients with cancer, yet they were prioritized in vaccination programs due to the higher risk of adverse outcomes (8–10). There is emerging evidence regarding safety and efficacy of vaccines in this cohort (11–13).

Vaccine hesitancy, defined as “a delay in acceptance or refusal of vaccination despite availability of vaccination services,” (14) has long been recognized as an important issue impacting vaccine uptake and the control of vaccine-preventable diseases (15). Indeed, vaccine hesitancy was declared one of the top threats to global health by the World Health Organization in 2019 (16). Vaccine hesitancy is a complex phenomenon, influenced by socio-demographic and attitudinal factors including age, gender, education, socio-economic status, beliefs about vaccines and trust in institutions (17).

Globally, numerous studies report differing rates of willingness to receive COVID-19 vaccines, including in people with underlying health conditions (18,19). Amongst people with cancer, vaccine hesitancy is generally lower than in the general population (19–21), although the decision-making impact of a cancer diagnosis on vaccination needs furtherexploration. People with cancer have expressed a range of concerns involving the impact of vaccines on the patients’ underlying cancer and treatments, as well as the potential effect of anti-cancer treatments on vaccine efficacy (22–25). Concern regarding vaccine-associated side effects and the potential to interfere with treatment schedules is also prominent (22,24,26–28). However, these cancer-specific factors and their influence on decision-making have not been comprehensively analyzed using validated assessment tools to assess vaccine hesitancy in the context of cancer.

To better understand vaccine hesitancy regarding COVID-19 vaccination in people with cancer, we conducted a large multicenter survey exploring how the cancer-specific context impacted vaccine attitudes and behavior. The use of three validated vaccine hesitancy scales allowed detailed understanding of factors influencing vaccination attitudes and the decision-making process.

## Materials and Methods

### Participants and Study Procedures

An internet-based cross-sectional survey hosted on the secure data capture platform Qualtrics (29) was conducted from 30 June to 06 October 2021. Eligible participants were aged 18 or over, with a past or current diagnosis of a solid organ or hematological malignancy, diabetes or multiple sclerosis (20). In this publication, we report on data from participants with a past or current diagnosis of a solid organ malignancy. Participants were recruited through nine Australian health services across four states, encompassing both public and private services in metropolitan and regional locations. The invitation to participate was sent by short message service to all patients with oncology clinic appointments in the various health services scheduled within the next six months and contained a link to the survey. Patients were also invited by clinicians during consultations and promotional materials were displayed at the health services. Tablet devices and paper surveys were available at some sites. After providing informed consent, participants completed eligibility questions and were directed to the survey. The survey was presented in English. The survey completion time was approximately 10 to 15 minutes. This study was approved by the Monash Health Human Research Ethics Committee (HREC) and the local research governance office at each recruitment site.

### Scales and Measures

The survey consisted of 42 items, including vaccine status, socio-demographics, cancer-related clinical details, as well as questions from three validated scales (Table S1):

1. Oxford COVID-19 Vaccine Hesitancy Scale (OHS), a 7-item scale measuring intent to receive a COVID-19 vaccine and vaccine hesitancy (30).
2. Oxford COVID-19 Vaccine Confidence and Complacency Scale (OCCS), a 14-item scale measuring attitudes around vaccine complacency and confidence. There are four identified factors: collective importance of a vaccine, belief that the vaccine will work, speed of vaccine development, and side-effects (30).
3. Disease Influenced Vaccine Acceptance Scale-Six (DIVAS-6), a 6-item scale evaluating the impact of cancer on participants’ attitudes towards COVID-19 vaccination (31). This scale was developed by the study team and validated in people with serious underlying medical conditions including cancer, diabetes, and multiple sclerosis. It consists of two subscales: the Disease-Complacency subscale, which assesses the degree to which a participant’s disease affected their perceived risk of COVID-19 infection, and the Vaccine-Vulnerability subscale, which assesses how the participant’s cancer diagnosis and treatment affected their perceived benefits and risks of the vaccine.

Consistent with the instrument’s instructions, responses to items on all three scales are recorded on a 5-point Likert scale, with an additional “Don’t Know” option provided. Higher scores indicate a greater degree of negative attitude towards vaccination. Responses for three DIVAS-6 items (questions 26, 27, 28) are reversed to be consistent with this pattern (higher scores indicating more negative attitude).

### Statistical Analysis

Summary and subscale scores for the OHS, OCCS and DIVAS-6 were calculated. ‘Don’t know’ responses were not analyzed, consistent with the scales’ methodology (30,32–34). Data cleaning was performed to remove unsubmitted incomplete, du-plicate, and ineligible responses. Missing data were not imputed.

Socio-demographic, clinical data, summary, and subscale scores were summarized using descriptive statistics. Variables with too few classifications were combined or removed on an analysis-by-analysis basis: this included combining no formal education level and primary education level and removing non-binary/other gender due to the small number of observations. Logistic regression was used to examine relationships between demographic and clinical factors and vaccination status (defined as having received one or more doses versus none). Linear regression was used to explore associations between demographic and clinical factors with scale summary and subscale scores. One-way ANCOVA was used to assess whether there were differences in scale summary and subscale scores between vaccination status. Time since study commencement was controlled in regression and ANCOVA to account for variation due to changing environmental circumstances. Hierarchical multivariable regression was conducted using demographic and disease-related variables that were significantly correlated at r >.10 with the outcome variable of interest (using Pearson’s and Spearman’s Rho). Cancer types were assessed as dichotomous variables compared to all other cancer types. Cancer types that showed a significant relationship with scale scores or vaccination status were assessed using hierarchical regression, controlling for significantly correlated (r>.10) demographic and disease-related variables. Alpha values of <0.05 were considered significant. Statistical analyses were conducted using SPSS Statistics Version 26.0 (IBM, USA).

## Results

### Patient Characteristics

Completed survey responses were received from 2691 people with solid organ cancers. Respondents had a median age of 62.5 years (*SD*=11.8; range 19-95), 40.9% were male, 71.3% lived in metropolitan areas, and 90.3% spoke English as their first language (Table 1). The most common cancer types were breast (36.6%), genitourinary (18.6%) and gastrointestinal (18.3%); the cancer was localized in 59.2% and 56.0% reported being currently on anti-cancer treatment.

**Table 1.** Participant characteristics

|  | **Total**<br>**n=2691 (%)** | **Vaccinated**<br>**n=2143 (79.7%)** | **Unvaccinated**<br>**n=546 (20.3%)** |
| --- | --- | --- | --- |
| Male | 1101 (40.9) | 898 (81.7) | 201 (18.3) |
| Female* | 1578 (58.6) | 1235 (78.3) | 343 (21.7) |
| Age: mean (SD) | 62.5 (11.8) | 63.7 (11.5) | 58.0 (12.0) |
| **Age (years)** |  |  |  |
| 18–49 | 396 (14.7) | 270 (68.2) | 126 (31.8) |
| 50–69 | 1444 (53.8) | 1113 (77.1) | 331 (22.9) |
| ≥70 | 847 (31.5) | 757 (89.5) | 89 (10.5) |
| **Highest level of education\*\*** |  |  |  |
| No formal/primary school | 72 (2.7) | 52 (72.2) | 20 (27.8) |
| Secondary school | 921 (34.3) | 726 (78.8) | 195 (21.2) |
| Vocational/Trade | 679 (25.2) | 526 (77.5) | 153 (22.5) |
| University | 1009 (37.5) | 834 (82.7) | 175 (17.3) |
| **Annual household Income (AUD)** |  |  |  |
| <50K | 898 (33.4) | 694 (77.3) | 204 (22.7) |
| 50K–100K | 653 (24.3) | 543 (83.2) | 110 (16.8) |
| 100K–150K | 364 (13.5) | 286 (78.6) | 78 (21.4) |
| >150K | 307 (11.4) | 269 (87.6) | 38 (12.4) |
| Prefer not to say | 467 (17.4) | 351 (75.2) | 116 (24.8) |
| **Aboriginal / Torres Strait Islander\*\*\*** |  |  |  |
| Yes | 38 (1.4) | 32 (84.2) | 6 (15.8) |
| **English as first language** |  |  |  |
| Yes | 2429 (90.3) | 1963 (80.9) | 464 (19.1) |
| No | 261 (9.7) | 180 (69.0) | 81 (31.0) |
| **Location** |  |  |  |
| Metropolitan | 1918 (71.3) | 1594 (83.1) | 324 (16.9) |
| Regional/rural | 773 (28.7) | 549 (71.2) | 222 (22.8) |
| **Cancer Type** |  |  |  |
| Breast | 986 (36.6) | 807 (81.8) | 179 (18.2) |
| Genitourinary | 501 (18.6) | 424 (84.8) | 76 (15.2) |
| Gastrointestinal | 493 (18.3) | 375 (76.1) | 118 (23.9) |
| Lung | 248 (9.2) | 194 (78.2) | 54 (21.8) |
| Skin | 151 (5.6) | 123 (82.0) | 27 (18.0) |
| Gynecological | 134 (5.0) | 91 (67.9) | 43 (32.1) |
| Head and Neck | 99 (3.7) | 69 (69.1) | 30 (30.3) |
| Other | 79 (2.9) | 60 (75.9) | 19 (24.1) |
| **Cancer Stage** |  |  |  |
| Localized | 1593 (59.2) | 1291 (81.1) | 301 (18.9) |
| Metastatic | 971 (36.1) | 770 (79.4) | 200 (20.6) |
| Unsure/Other | 127 (4.7) | 82 (64.6) | 45 (35.4) |
| **Time since diagnosis** |  |  |  |
| <6 months | 372 (13.8) | 251 (67.5) | 121 (32.5) |
| 6–24 months | 964 (35.8) | 755 (78.4) | 208 (21.6) |
| 2–5 years | 841 (31.3) | 695 (82.6) | 146 (17.4) |
| >5 years | 514 (19.1) | 442 (86.2) | 71 (13.8) |
| **Current anti-cancer treatment** |  |  |  |
| Yes | 1506 (56.0) | 1181 (78.5) | 323 (21.5) |
| No | 1185 (44.0) | 962 (81.2) | 223 (18.8) |

**Notes:** Exact cohort for vaccination status n =2689. \*does not include "non-binary/prefer not to say" n=12 (0.4%) \*\*Other: 8 (0.3%); \*\*\*does not include "prefer not to say" n=27 (1.0%). Abbreviations: AUD, Australian Dollars; K, 1000.

### COVID-19 Vaccine Uptake

The main features of the Australian vaccine rollout program and its relation to community transmission of COVID-19 and other public health measures are shown in Figure 1, giving context to the health and social environment at the time of the survey. The vaccination rate for the total study population was 79.7%. Female gender (OR 0.77, 95% CI 0.63, 0.95, p = 0.012), English as the non-dominant language (OR 0.69, 95% CI 0.51, 0.93, p = 0.02), and regional/rural location (OR 0.65, 95% CI 0.53, 0.80, p < 0.001) were associated with a significantly decreased likelihood of vaccine uptake (Figure 2 and Table S2). Conversely, older age (50-69 years [OR 1.53, 95% CI 1.18, 1.99, p = 0.002) and annual household income $50 000-$100 000 Australian Dollars (AUD; OR 1.35, 95% CI 1.03, 1.76, p = 0.03) were associated with significantly higher likelihood of being vaccinated.

**Figure 1.**
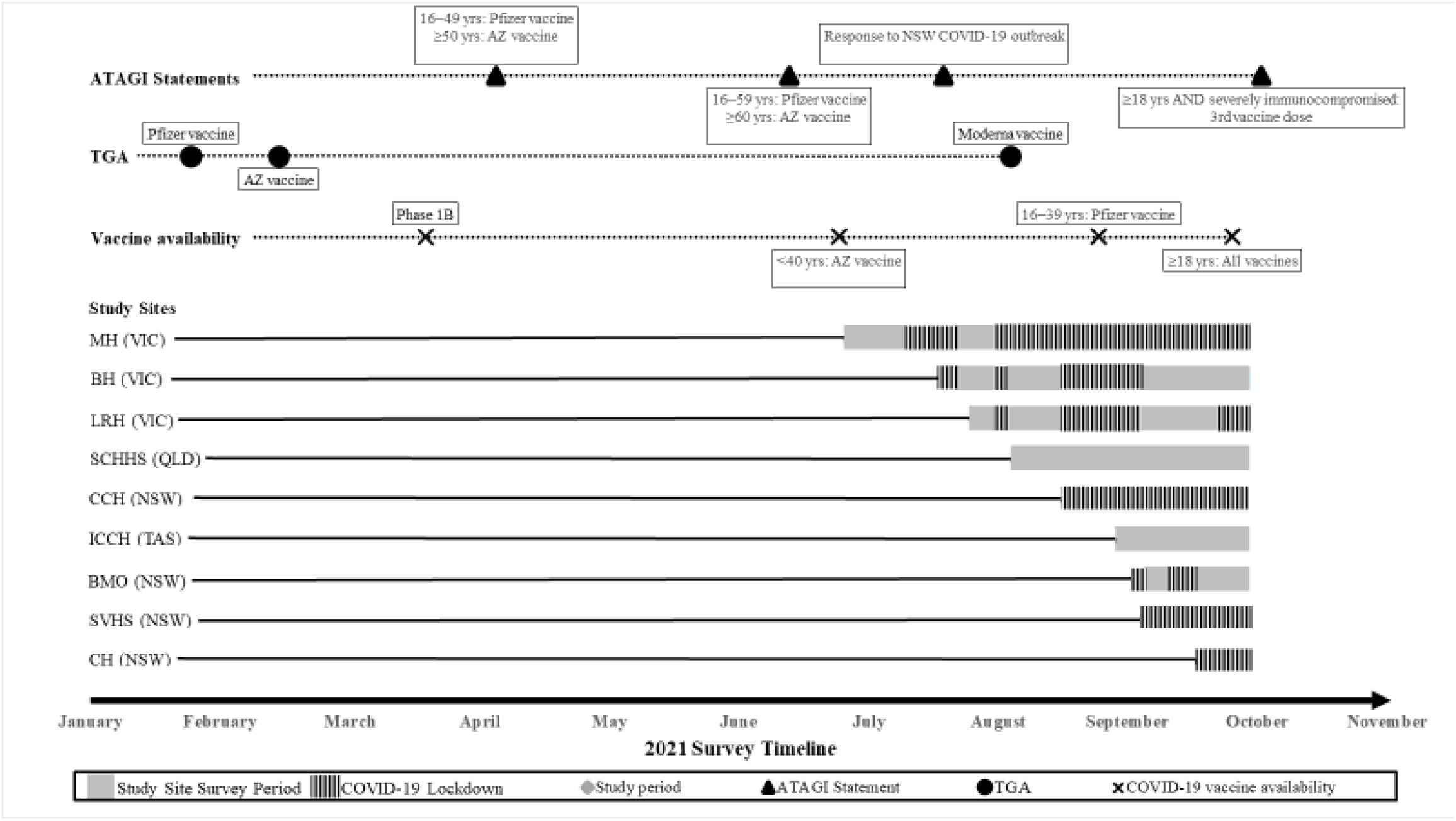
Timeline of study including survey period relative to state-wide strict lockdowns for COVID -19. Abbreviations: ATAGI, Australian Technical Advisory Group on Immunization; Years, yrs; AZ, Astra-Zeneca; NSW, New South Wales; TGA, Therapeutic Goods Administration – COVID-19 Vaccine Provisional Registration; MH, Monash Health; VIC, Victoria; BH, Bendigo Health; LRH, Latrobe Regional Hospital; SCHHS; Sunshine Coast Hospital and Health Service; QLD; Queensland; CCH, Central Coast Hematology; ICCH, Icon Cancer Center Hobart; TAS, Tasmania; BMO, Border Medical Oncology; SVHS, St Vincent’s Hospital Sydney; CH, Campbelltown Hospital. People with solid organ tumors were eligible for COVID-19 vaccination from the commencement of the Australian Government Rollout Phase 1B.

**Figure 2.**
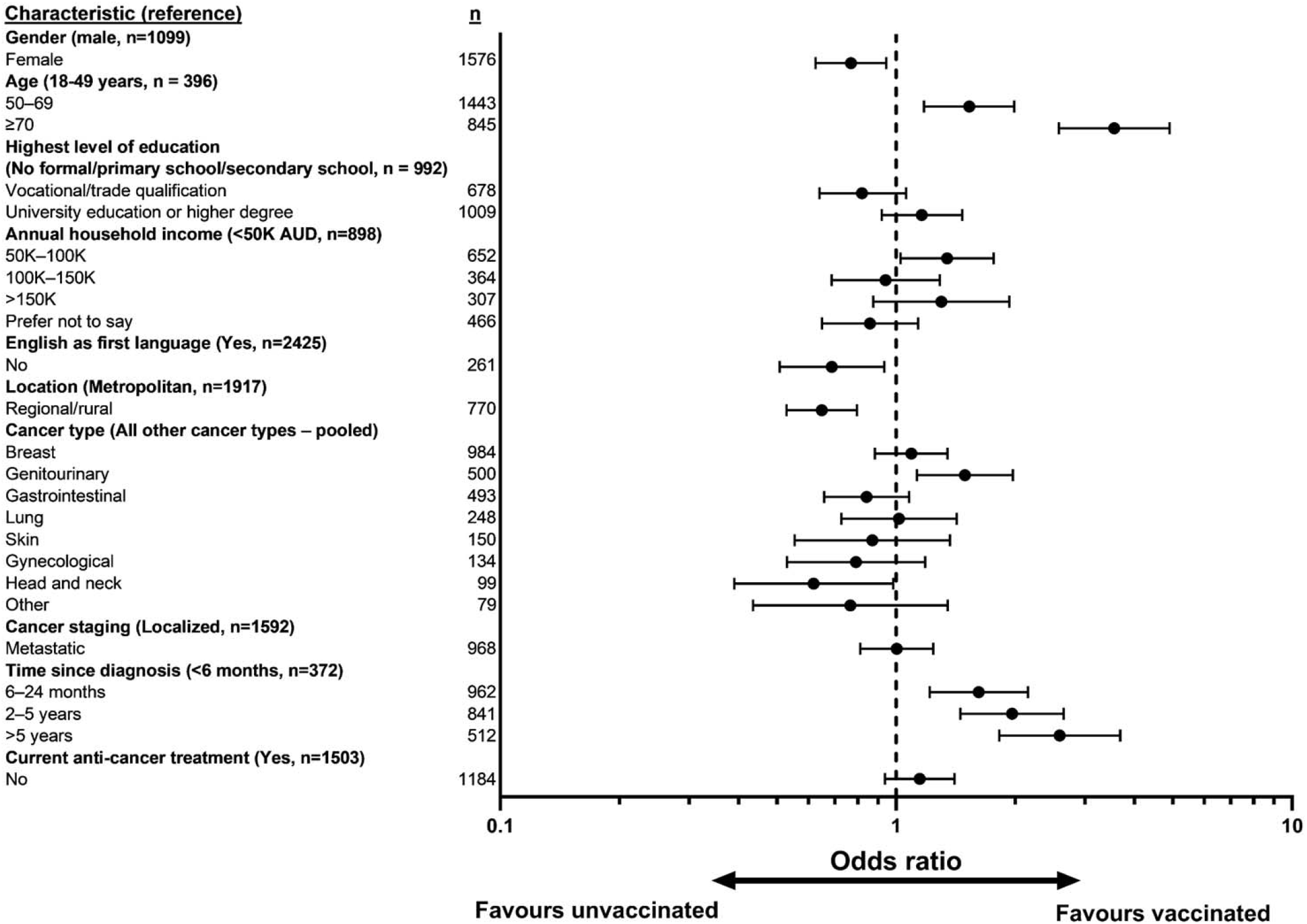
Forest plot of time-adjusted logistic regression model of factors associated with vaccine uptake. Abbreviations: AUD, Australian dollars; K, 1000.

With regard to cancer characteristics (Figure 2 and Table S2), time-adjusted analysis showed higher vaccination rate amongst patients with a longer time since cancer diagnosis (6-24 months [OR 1.62, 95% CI 1.22, 2.15, p<0.001], 2-5 years [OR 1.96, 95% CI 1.45, 2.65, p<0.001], >5 years [OR 2.59, 95% CI 1.82, 3.68, p<0.001]) and with the primary site in the genitourinary system (OR 1.49, 95% CI 1.13, 1.97, p = 0.005). People with head and neck cancer had a significantly lower likelihood of vaccine uptake (OR 0.62, 95% CI 0.39, 0.98, p = 0.04).

On multivariable analysis, age, regional location, and time since cancer diagnosis remained significant predictors of vaccine status (Table S3). The relationship of being vaccinated with genitourinary cancer type was no longer significant after controlling for age, gender, location, and time since cancer diagnosis (Table S4). The converse relationship with head and neck cancer remained significant after controlling for the same factors (Table S5).

### Oxford COVID-19 Vaccine Hesitancy Scale

Significantly higher Oxford COVID-19 Vaccine Hesitancy Scale summary scores, indicating greater vaccine hesitancy, were observed in unvaccinated respondents, (adjusted mean difference 7.38 [95% CI 6.93, 7.84, p <0.001]).

When independently analyzed, higher hesitancy scores were significantly associated with female gender; younger age and English as the non-dominant language (Table S6). When entered together using multivariate analysis, age, university education and English as a second language remained significant (Table S7). Genitourinary cancer was associated with lower vaccine hesitancy scores, and this remained significant after adjusting for relevant sociodemographic and clinical variables (Table S8).

### Oxford COVID-19 Vaccine Confidence and Complacency Scale

Higher scores, indicating more prevalent concerns surrounding vaccine efficacy and side effects, were observed in unvaccinated participants. No significant demographic or cancer-related associations were seen.

### Disease Influenced Vaccine Acceptance Scale-6

Significantly higher summary and subscale scores were observed in unvaccinated participants, who reported more concerns about vaccine efficacy (60.2% v. 34.2%), side-effects (72.1% v. 28.9%) and interactions with anti-cancer treatment (53.4% v. 17.7%) (Figure 3a). Regarding the statement ‘cancer makes me more worried about being infected with COVID-19,’ 56.8% agreed (comprising strongly agree or somewhat agree); this was independent of vaccination status (Figure 3b). The statement ‘my cancer means having the vaccine is more important to me’ was agreed with by 67.2% overall (72.3% of vaccinated and 45.7% of unvaccinated participants). Vaccinated respondents were more likely to agree with the importance of a doctor’s recommendation regarding the vaccine (81.7% v. 66.0%).

**Figure 3.**
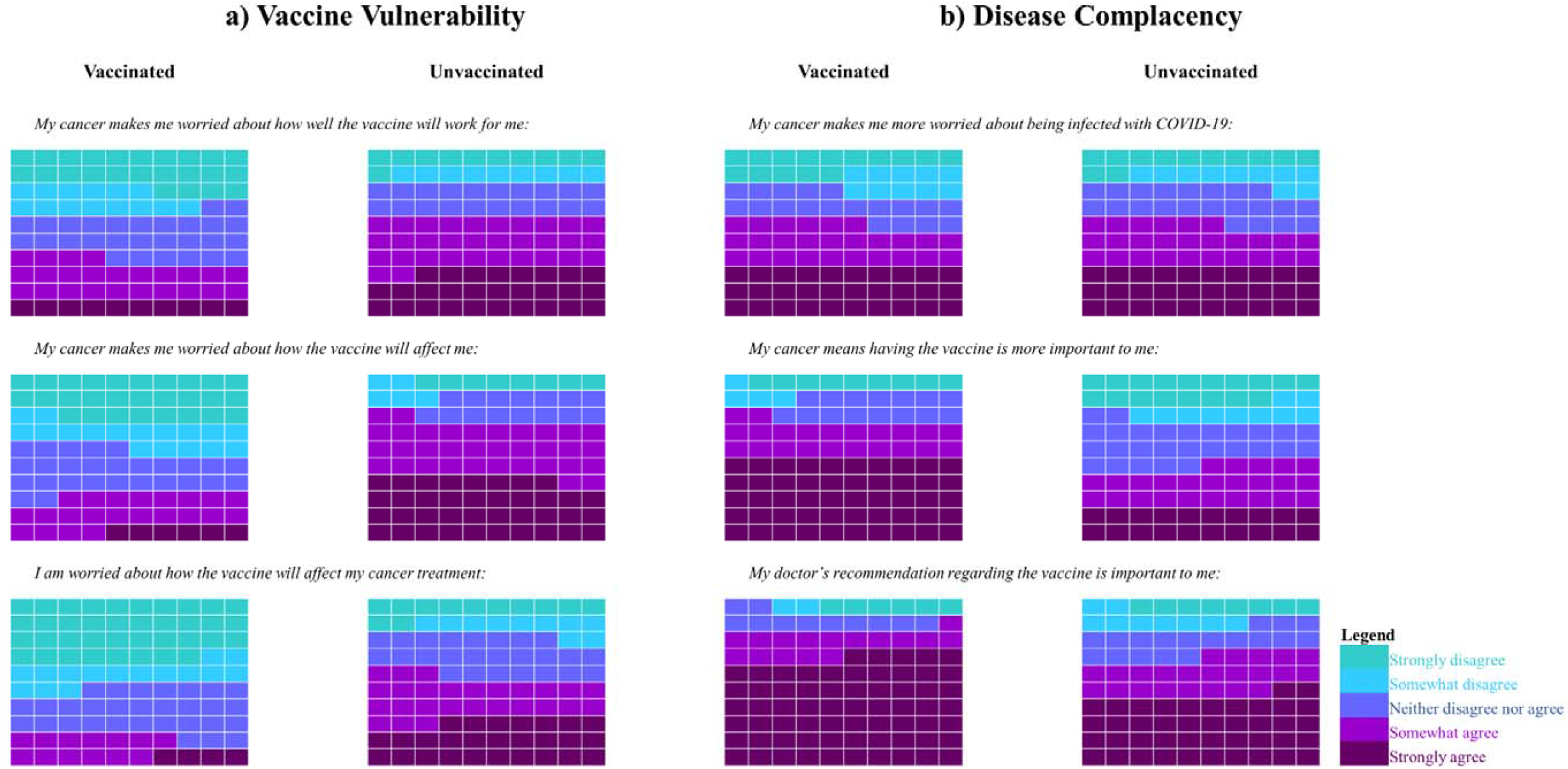
Response frequencies for the DIVAS-6 subscales: a) Vaccine Vulnerability b) Disease Complacency. Each box represents 1% of responses. Abbreviations: DIVAS-6, Disease Influenced Vaccine Acceptance Scale-6.

Analysis of DIVAS-6 subscale scores across demographic and cancer characteristics identified differences in disease complacency and vaccine vulnerability scores (Figure 4, Tables S9 and S10). Participants with metastatic cancer stage, on active cancer treatment or with lung cancer reported significantly higher vaccine vulnerability coupled with significantly lower disease complacency scores, identifying participants who had greater concerns about contracting COVID-19 due to their cancer coupled with greater concerns about the effect of the vaccine on disease progression and treatments. Conversely, those with localized cancer stage and not on active treatment or with genitourinary cancer reported significantly lower vaccine vulnerability and higher disease complacency, identifying participants who were less concerned with contracting COVID-19 due to their cancer coupled with low vaccine-related concerns.

**Figure 4:**
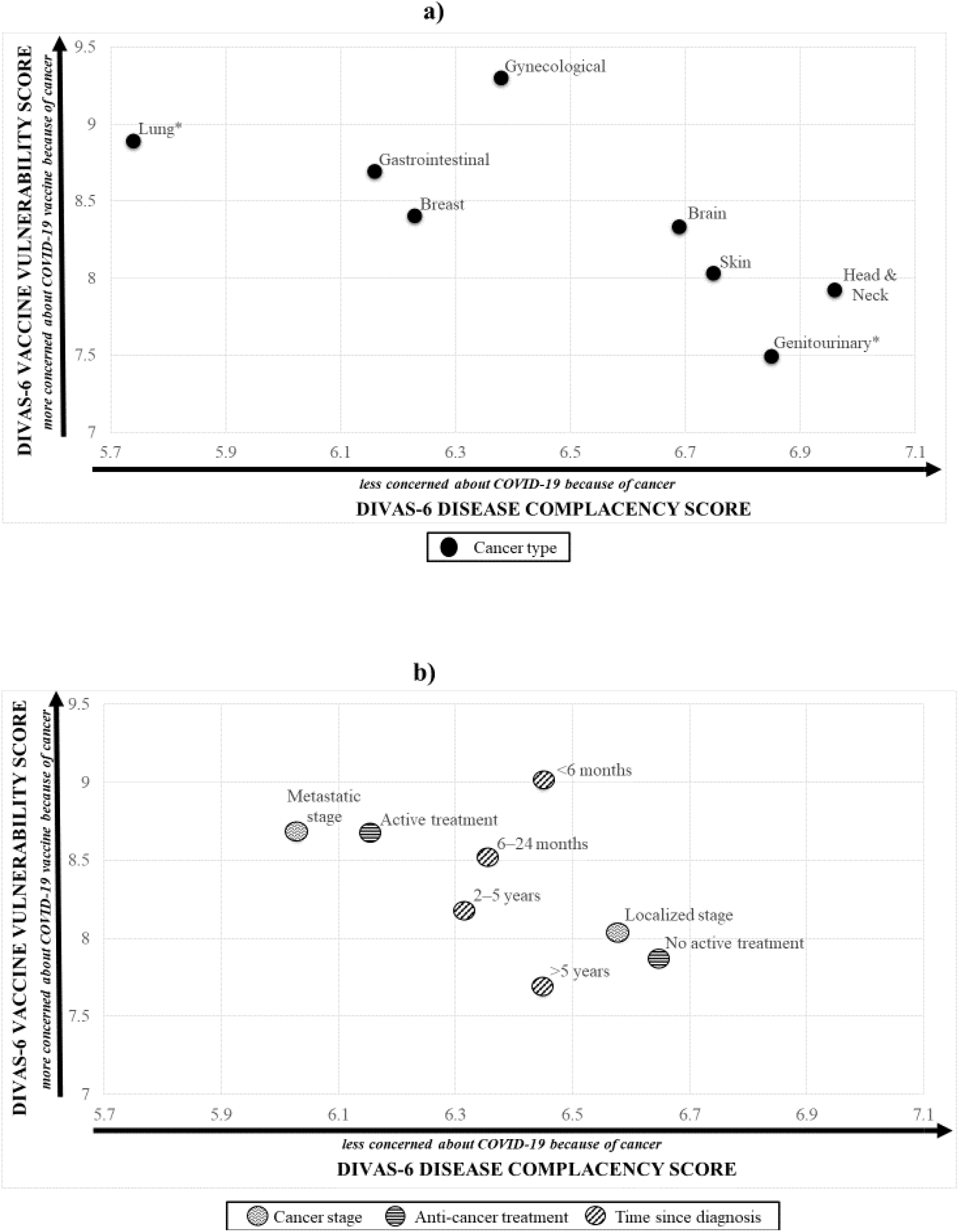
DIVAS-6 Disease Complacency against Vaccine Vulnerability scores by a) cancer type b) clinical characteristics. Abbreviations: DIVAS-6, Disease Influenced Vaccine Acceptance Scale-6.

## Discussion

This study comprehensively assessed COVID-19 vaccine hesitancy specifically in people with cancer using a suite of validated scales, including a disease-specific scale to assess the influence of having cancer on vaccine attitudes. Over half of our large and diverse sample expressed a perception of vulnerability regarding COVID-19 infection due to their underlying cancer, and this was irrespective of vaccination status. Concurrently, there were prominent concerns regarding the impact of cancer on vaccine efficacy, adverse effects, and interactions with anti-cancer treatments; this was particularly strong amongst unvaccinated participants.

Interestingly, the overall vaccination rate (79.7%) was not higher than the general Australian population at the time (80.5%), despite people with cancer having two extra months access to free vaccines (35). This finding potentially reflects the influence of the disease-specific factors faced by people with cancer, adding additional concerns to this complex decision-making process. Consistent with smaller studies, there were few differences in uptake and hesitancy between people with different cancer types and stages or treatment status (23,24,36,37). In comparison to the original study conducted in a UK representative general population group, our respondents displayed a much lower mean score on the Oxford COVID-19 Vaccine Hesitancy Scale. This likely reflects the more medically vulnerable population in our study, the impact of having effective vaccines available and the reduction in vaccine hesitancy over the course of pandemic, a trend shown in a number of studies (30,38). Despite this, significant concerns regarding vaccine efficacy and safety were evident as demonstrated by the present study, highlighting the ongoing need to address these concerns as the pandemic continues, especially given the recommendations for booster and additional vaccine doses in cancer populations (39–41).

The DIVAS-6 scale provides additional insight into how a cancer diagnosis impacts both the fear of COVID-19 disease and any hesitancy toward vaccination (and conversely, acceptance). It appears to be a useful addition to the two Oxford scales, which largely examine attitudes and beliefs independent of other factors affecting a person’s health. For example, increased hesitancy was observed in people diagnosed within 6 months and those with metastatic disease. This is consistent with previous studies which suggest that these groups would likely be susceptible to increased cancer-related anxiety due to increased uncertainty surrounding their conditions (42). It follows that such patient groups, once identified, can be more specifically targeted by public information campaigns, as well as alerting individual clinicians that a discussion about how COVID-19 and vaccination may affect a particular patient may be required. Our findings suggest that patients from culturally and linguistically diverse backgrounds and those living in regional/rural areas would benefit from such targeted strategies.

Our survey reinforces previous studies showing that healthcare professionals are a trusted resource for patients with cancer (26,43,44). In a Korean study of 1001 people with cancer, 61% indicated they were willing to accept a vaccine when available; this increased to 91% with a recommendation from their physician (37). Furthermore, a physician-led educational intervention in the form of a webinar was demonstrated to increase patient understanding about vaccines and increase their intention to be vaccinated (45). As such, clinicians should be harnessed as an important partner in vaccination campaigns, which is particularly relevant now it appears that ongoing boosters against SARS-CoV-2 will be required.

Numerous professional bodies have produced guidelines and educational resources for patients and clinicians. At the start of the pandemic these were based on expert opinion, but now need to evolve to incorporate emerging data about vaccine efficacy and timing, tailored to different patient cohorts (46–49). In addition to the challenge of keeping standard but tailored culturally appropriate information up-to-date, focus on harmonized dissemination is required to ensure consistency of advice is provided by the many other practitioners encountered by people with cancer, as well as with public education campaigns(50). We have previously documented the negative impact of receiving discordant information for people with cancer (27,51).

Insights provided by DIVAS-6 suggest it is a useful tool to facilitate individualized discussions between clinicians and patients with cancer. The two subscales elucidate the influences underlying a person’s attitude towards vaccination; whether having a cancer diagnosis raised perceived vulnerabilities to COVID-19 infection and therefore was a source of motivation for vaccination, or the contrary where the main concerns were that vaccines could add risks to the person’s cancer or treatments. The six questions can be easily implemented in clinical environments and attitudes tracked over time, which is important given our data showing hesitancy reduced with longer time since diagnosis.

As with any survey, there are some limitations, including representation of those who chose to complete the survey and recall and misclassification bias. Our study population had a slightly greater proportion of females, likely related to the high proportion of participants with breast cancer. We did not evaluate broader cultural and political factors which have been found to significantly affect attitudes towards vaccination such as social media usage, socio-political group association, or adherence to COVID-19 vaccine conspiracy theories but rather focused on disease-related influences (52–54). Additionally, the epidemiology of COVID-19 in Australia at the time of the study, with comparably low prevalence and disease burden, may somewhat limit direct comparisons with other regions of the world; more likely, much of the information is likely to be valuable across many jurisdictions.

This is a large study of COVID-19 vaccine hesitancy in people with cancer, sampling across a diverse range of demographics and cancer-disease characteristics in Australia. The use of multiple validated tools, including disease-specific measures, has given a comprehensive picture of attitudes of this medically vulnerable group and can be used to target education to further raise vaccine compliance.

## Conclusion

In summary, vaccine attitudes and hesitancy in people with cancer are substantially influenced by their underlying cancer diagnosis. Medical vulnerability to COVID-19 due to disease status or anti-cancer treatments can motivate and enable positive vaccine uptake; conversely, concerns regarding a potential negative impact of the vaccine on health, underlying cancer and treatments contributes to hesitancy. These complex concerns must be comprehensively addressed both in individual patients and in public health messaging, to best protect this medically vulnerable population from the ongoing morbidity and mortality of the COVID-19 pandemic.

## Data Availability

All data produced in the present study are available upon reasonable request to the authors.

## Acknowledgements

We thank our consumer representative Janne Williams, all study participants, and the recruitment teams at each study site.

## Data availability statement

All data produced in the present study are available upon reasonable request to the authors.

