## Supplementary materials for "COVID-19 vaccine hesitancy in Australian patients with solid organ cancers"

**CANcer patients' perspectives on coronavirus VACCination Survey (CANVACCS) investigators:**
*Monash Health, Victoria*: Dr. Veronica Lopez Aedo, Dr. Elizabeth Ahern, Dr. Muhammad Alamgeer, Dr. Nathan Bain, Dr. Amy Body, A/Prof. Peter Briggs, Dr. Daphne Day, Dr. Sophia Frentzas, Dr. Lisa Grech, A/Prof. Marion Harris, Dr. Gwo-Yaw Ho, Dr. Caroline Lum, Dr. Vi Luong, Dr. Amelia McCartney, Dr. Cameron McLaren, Dr. Mike Nguyen, Prof. Stephen Opat, A/Prof. David Pook, Prof. Eva Segelov, A/Prof. Andrew Strickland, Dr. Avraham Travers, Dr. Kate Webber, Dr. Michelle White, Dr. Walid Zwieky.
*Bendigo Health, Victoria*: Dr. Sam Harris.
*Latrobe Regional Hospital, Victoria:* Dr. Hieu Chau.
*Sunshine Coast Hospital and Health Service, Queensland:* A/Prof. Bryan Chan.
*Icon Cancer Centre Hobart, Tasmania:* A/Prof. Louise Nott.
*Central Coast Hematology, New South Wales:* Dr. Richard Blennerhassett, Dr. Cecily Forsyth, Ms. Jacqueline Jagger.
*St Vincent's Hospital Sydney, New South Wales:* A/Prof. Nada Hamad.
*Campbelltown Hospital, New South Wales:* Dr. Annette Tognela.
*Border Medical Oncology, New South Wales:* A/Prof. Craig Underhill.

**Table S1.** Online survey questions

| | |
|---|---|
| **Screening items** | |
| Are you 18 years or older? | o Yes<br>o No (Terminate if No) |
| Have you received a cancer diagnosis? | o Yes<br>o No (Terminate if No) |
| Are you a [participating site] patient? | o Yes<br>o No (Terminate if No) |
| **Vaccination status** | |
| Have you already received a COVID-19 vaccine? | o Yes, 1 dose only<br>o Yes, 2 doses<br>o No |
| **Oxford COVID-19 Vaccine Hesitancy Scale**<br>Instructions: We would like to know your feelings and thoughts about the COVID-19 vaccine. Note: If you have already been vaccinated against COVID-19, please complete these questions in relation to a future COVID-19 vaccine dose/booster. | |
| Would you take a COVID-19 vaccine if offered? | o Definitely/have taken<br>o Probably<br>o I may or may not<br>o Probably not<br>o Definitely not<br>o Don't know |
| When a COVID-19 vaccine is available: | o I will want to get it as soon as possible<br>o I will take it when offered<br>o I'm not sure what I will do<br>o I will put off (delay) getting it<br>o I will refuse to get it<br>o Don't know |
| I would describe my attitude towards receiving a COVID-19 vaccine as: | o Very keen<br>o Pretty positive<br>o Neutral<br>o Quite uneasy<br>o Against it<br>o Don't know |
| If a COVD-19 vaccine was available in my local area, I would: | o Get it as soon as possible<br>o Get it when I have time<br>o Delay getting it<br>o Avoid getting it for as long as possible<br>o Never get it<br>o Don't know |
| If my family or friends were thinking of getting a COVID-19 vaccination, I would: | o Strongly encourage them<br>o Encourage them<br>o Not say anything to them about it<br>o Ask them to delay getting the vaccination<br>o Suggest that they do not get the vaccination<br>o Don't know |
| I would describe myself as: | o Eager to get a COVID-19 vaccine<br>o Willing to get the COVID-19 vaccine<br>o Not bothered about getting the COVID-19 vaccine<br>o Unwilling to get the COVID-19 vaccine<br>o Anti-vaccination for COVID-19<br>o Don't know |
| Taking a COVID-19 vaccination is: | o Really important<br>o Important<br>o Neither important nor unimportant<br>o Unimportant<br>o Really unimportant<br>o Don't know |
| Oxford COVID-19 Vaccine Confidence and Complacency Scale | |
| Do you think you will be infected with COVID-19 over the next 12 months? | o Definitely<br>o Probably<br>o Possibly |

| | |
|---|---|
| | o Probably not<br>o Definitely not<br>o Don’t know |
| I think the COVID-19 vaccine is likely to: | o Work for almost everyone<br>o Work for most people<br>o I am unsure how many people it will work for<br>o Not work for most people<br>o Not work for anyone<br>o Don’t know |
| I think the COVID-19 vaccine is likely to: | o Definitely work for me<br>o Probably work for me<br>o May or may not work for me<br>o Probably not work for me<br>o Definitely not work for me<br>o Don’t know |
| I think if I get the COVID-19 vaccine it will be: | o Really helpful for the community around me<br>o Helpful for the community around me<br>o Neither helpful nor unhelpful for the community around me<br>o Unhelpful for the community around me<br>o Really unhelpful for the community around me<br>o Don’t know |
| I think if individuals like me get the COVID-19 vaccine it will: | o Save a large number of lives<br>o Save some lives<br>o Have no impact<br>o Lead to more deaths<br>o Lead to a large number of deaths<br>o Don’t know |
| I think the speed of developing and testing the vaccine means it will be: | o Really good<br>o Good<br>o Will not affect how good or bad it is<br>o Bad<br>o Really bad<br>o Don’t know |
| I think the speed of developing and testing the vaccine means it will be: | o Really safe<br>o Safe<br>o It will not affect how safe it is<br>o Unsafe<br>o Really unsafe<br>o Don’t know |
| I think if many people do not get the vaccine this: | o Will be dangerous<br>o May be dangerous<br>o Will have no consequences at all<br>o May be good<br>o Will be good<br>o Don’t know |
| I expect that receiving the vaccine will be: | o Hardly noticeable<br>o A little unpleasant<br>o Moderately unpleasant<br>o Painful<br>o Extremely painful<br>o Don’t know |
| I think the side-effects for people of getting the COVID-19 vaccine will be: | o None<br>o Mild<br>o Moderate<br>o Significant<br>o Life-threatening<br>o Don’t know |
| I think the COVID-19 vaccine will: | o Greatly strengthen my immune system<br>o Strengthen my immune system<br>o It will neither strengthen nor weaken my immune system<br>o Weaken my immune system<br>o Greatly weaken my immune system<br>o Don’t know |

| | |
|---|---|
| I think taking the COVID-19 vaccine: | o Will give me complete freedom to get on with life just as before<br>o Will give me greater freedom<br>o Will have no effect on my freedom<br>o Will restrict my freedom<br>o Will completely restrict my freedom to get on with life<br>o Don't know |
| I think getting the vaccine is a sign of: | o Great personal strength<br>o Personal strength<br>o Not a sign of personal strength or weakness<br>o Personal weakness<br>o Great personal weakness<br>o Don't know |
| Taking a new COVID-19 vaccine will make me feel like a guinea pig: | o Do not agree<br>o Agree a little<br>o Agree moderately<br>o Agree a lot<br>o Completely agree<br>o Don't know |
| **Disease Influenced Vaccine Acceptance Scale-Six**<br><br>Instructions: We would like to know about how your cancer diagnosis may be related to your feelings and thoughts about the COVID-19 vaccine. For each of the following statements, please tap/click the one choice that best represents how strongly you agree or disagree with it. There are 6 choices to choose from for each statement. | |
| My history of cancer makes me more worried about being infected with COVID-19: | o Strongly agree<br>o Somewhat agree<br>o Neither disagree nor agree<br>o Somewhat disagree<br>o Strongly disagree<br>o Don't know |
| My history of cancer means having the vaccine is more important to me: | o Strongly agree<br>o Somewhat agree<br>o Neither disagree nor agree<br>o Somewhat disagree<br>o Strongly disagree<br>o Don't know |
| My doctor's recommendation regarding the vaccine is important to me: | o Strongly agree<br>o Somewhat agree<br>o Neither disagree nor agree<br>o Somewhat disagree<br>o Strongly disagree<br>o Don't know |
| My history of cancer makes me worried about how well the vaccine will work for me: | o Strongly disagree<br>o Somewhat disagree<br>o Neither disagree nor agree<br>o Somewhat agree<br>o Strongly agree<br>o Don't know |
| My history of cancer makes me worried about how the vaccine will affect me: | o Strongly disagree<br>o Somewhat disagree<br>o Neither disagree nor agree<br>o Somewhat agree<br>o Strongly agree<br>o Don't know |
| I am worried about how the vaccine will affect my cancer treatment: | o Strongly disagree<br>o Somewhat disagree<br>o Neither disagree nor agree<br>o Somewhat agree<br>o Strongly agree<br>o Don't know |
| **Clinical** | |
| What type of cancer do you have? (i.e. where your cancer started and not where it has spread to). | o Breast<br>o Lung (including mesothelioma)<br>o Genitourinary (i.e. prostate, kidney, testicular or bladder)<br>o Skin (including melanoma) |

| | |
|---|---|
| | o Gastrointestinal (i.e. stomach, esophagus, bile duct, gallbladder, pancreas, colon, rectum, or anus)<br>o Gynecological (i.e. ovarian, cervical, uterine or vulvar/vaginal)<br>o Head and neck (i.e. mouth, throat, sinus or nose)<br>o Brain<br>o Blood (i.e. leukemia, myeloma and lymphoma)<br>o Other (please specify):___________________________ |
| When was your cancer diagnosed? | o Less than 6 months ago<br>o 6 to 24 months ago<br>o 2 to 5 years ago<br>o More than 5 years ago |
| As you understand it, is your cancer in just the one area where it started (localized) or has it spread to other places in the body (metastatic)? | o Localized<br>o Metastatic<br>o Don't know<br>o Other (please type any comments): _____________________ |
| Are you currently on cancer treatment? | o Yes<br>o No |
| How long ago was your last treatment? (e.g. chemotherapy, immunotherapy, hormonal treatment, targeted therapy, radiotherapy and/or clinical trial). | o Currently on treatment<br>o Less than 1 year ago<br>o 1 to 5 years ago<br>o More than 5 years ago |
| Socio-demographics | |
| What is your gender? | o Male<br>o Female<br>o Non-binary / third gender<br>o Prefer not to say |
| What is your age? | _________________ |
| What is your highest educational level (completed)? | o No formal education<br>o Primary education<br>o Secondary education<br>o Vocational/trade qualification<br>o University education or higher degree<br>o Other (please specify): _____________________ |
| What is your annual household income (including everyone who lives in your home)? | o Less than $50,000<br>o $50,001 to $100,000<br>o $100,001 to $150,000<br>o More than $150,000<br>o Prefer not to say |
| Do you identify as Aboriginal and/or Torres Strait Islander? | o Yes<br>o No<br>o Prefer not to say |
| Is English your first language? | o Yes<br>o No |
| Please include any comments about your feelings and thoughts about your cancer and COVID-19 vaccination that you would like to share. If you have no comments to include, please type 'Nil'. | [Text entry]: _______________________________________ |

**Table S2.** Logistic regression analysis predicting vaccinated status with sociodemographic and clinical characteristics.

| Category (reference) | n | B (SE) | Odds ratio (95% CI) | p-value |
|---|---|---|---|---|
| Gender (Male) | 2675 | | | |
| Female | | -0.26 (0.11) | 0.77 (0.63 – 0.95) | 0.012 |
| Age (18–49 years) | 2684 | | | |
| 50–69 | | 0.43 (0.13) | 1.53 (1.18 – 1.99) | 0.002 |
| ≥70 | | 1.27 (0.16) | 3.56 (2.58 – 4.91) | <0.001 |
| Highest level of education (No formal /primary school/secondary school) | 2679 | | | |
| Vocational/Trade | | -0.20 (0.13) | 0.82 (0.64 – 1.06) | 0.13 |
| University | | 0.15 (0.12) | 1.16 (0.92 – 1.47) | 0.21 |
| Annual household income (AUD<50K) | 2687 | | | |
| 50K–100K | | 0.30 (0.14) | 1.35 (1.03 – 1.76) | 0.03 |
| 100K–150K | | -0.06 (0.16) | 0.94 (0.69 – 1.29) | 0.70 |
| >150K | | 0.26 (0.20) | 1.30 (0.88 – 1.93) | 0.19 |
| Prefer not to say | | -0.15 (0.14) | 0.86 (0.65 – 1.14) | 0.29 |
| English as first language (Yes) | 2686 | | | |
| No | | -0.37 (0.16) | 0.69 (0.51 – 0.93) | 0.02 |
| Location (Metropolitan) | 2687 | | | |
| Regional/rural | | -0.43 (0.10) | 0.65 (0.53 – 0.80) | <0.001 |
| Cancer Type (All other cancer types – pooled) | 2687 | | | |
| Breast | | 0.09 (0.11) | 1.09 (0.88 – 1.35) | 0.42 |
| Genitourinary | | 0.40 (0.14) | 1.49 (1.13 – 1.97) | 0.005 |
| Gastrointestinal | | -0.17 (0.13) | 0.84 (0.66 – 1.08) | 0.17 |
| Lung | | 0.02 (0.17) | 1.02 (0.73 – 1.42) | 0.92 |
| Skin | | -0.14 (0.23) | 0.87 (0.55 – 1.37) | 0.55 |
| Gynecological | | -0.23 (0.21) | 0.79 (0.53 – 1.18) | 0.26 |
| Head and Neck | | -0.48 (0.24) | 0.62 (0.39 – 0.98) | 0.04 |
| Other | | -0.27 (0.29) | 0.77 (0.44 – 1.35) | 0.36 |
| Cancer Stage (Localized) | 2560 | | | |
| Metastatic | | 0.004 (0.11) | 1.00 (0.81 – 1.24) | 0.97 |
| Time since diagnosis (<6 months) | 2687 | | | |
| 6–24 months | | 0.48 (0.15) | 1.62 (1.22 – 2.15) | 0.001 |
| 2–5 years | | 0.67 (0.15) | 1.96 (1.45 – 2.65) | <0.001 |
| >5 years | | 0.95 (0.18) | 2.59 (1.82 – 3.68) | <0.001 |
| Current anti-cancer treatment (Yes) | 2687 | | | |
| No | | 0.14 (0.10) | 1.15 (0.94 – 1.40) | 0.18 |

**Notes:** Regression analyses was controlled for time since study commencement. These variables were excluded due to <50 responses; Aboriginal and/or Torres Strait Islander status: non-binary/other gender, and "other" educational level. Cancer stage (don't know/other) were excluded for comparisons between localized and metastatic. Abbreviations: B, unstandardized coefficient; SE, standard error; 95% CI, 95% confidence interval; AUD, Australian Dollars; K, 1000.

**Table S3.** Hierarchical multivariable logistic regression analysis of vaccinated status (n = 2684).

| | B | SE | Wald | df | p-value | OR | 95% CI | |
|---|---|---|---|---|---|---|---|---|
| | | | | | | | Lower | Upper |
| **Time since study commencement** | 0.03 | 0.002 | 205.10 | 1 | <0.001 | 1.03 | 1.03 | 1.04 |
| **Age (years)** | | | | | | | | |
| 18–49 (reference) | - | - | - | - | - | - | - | - |
| 50–69 | 0.44 | 0.14 | 10.39 | 1 | 0.001 | 1.55 | 1.19 | 2.02 |
| ≥70 | 1.27 | 0.17 | 58.07 | 1 | <0.001 | 3.56 | 2.57 | 4.94 |
| **Location** | | | | | | | | |
| Metropolitan (reference) | - | - | - | - | - | - | - | - |
| Regional/rural | -0.47 | 0.11 | 19.54 | 1 | <0.001 | 0.62 | 0.51 | 0.77 |
| Time since diagnosis | | | | | | | | |
| <6 months (reference) | - | - | - | - | - | - | - | - |
| 6–24 months | 0.50 | 0.145 | 11.29 | 1 | 0.001 | 1.65 | 1.23 | 2.20 |
| 2–5 years | 0.69 | 0.16 | 19.67 | 1 | <0.001 | 2.00 | 1.47 | 2.72 |
| >5 years | 0.83 | 0.18 | 20.43 | 1 | <0.001 | 2.29 | 1.60 | 3.28 |
| Constant | -1.03 | 0.19 | 30.61 | 1 | <0.001 | 0.36 | | |

**Notes:** Variables entered at each step: Step 1, days since study commencement; Step 2, age and location; Step 3, time since diagnosis. Abbreviations: B, unstandardized coefficient; SE, standard error; df, degrees of freedom; OR, odds ratio; 95% CI, 95% confidence interval; AUD, Australian Dollars; K, 1000.

**Table S4.** Hierarchical multivariable logistic regression analysis of vaccinated status with genitourinary cancer type, compared with all other cancer types (n = 2545).

| | B | SE | Wald | df | p-value | OR | 95% CI | |
|---|---|---|---|---|---|---|---|---|
| | | | | | | | Lower | Upper |
| **Time since study commencement** | 0.03 | 0.002 | 185.25 | 1 | <0.001 | 1.03 | 1.03 | 1.04 |
| **Genitourinary cancer type** | 0.06 | 0.18 | 0.11 | 1 | 0.74 | 1.06 | 0.75 | 1.50 |
| **Age (years)** | | | | | | | | |
| 18–49 (reference) | - | - | - | - | - | - | - | - |
| 50–69 | 0.46 | 0.14 | 10.37 | 1 | 0.001 | 1.58 | 1.20 | 2.08 |
| ≥70 | 1.27 | 0.18 | 50.77 | 1 | <0.001 | 3.57 | 2.51 | 5.06 |
| **Location** | | | | | | | | |
| Metropolitan (reference) | - | - | - | - | - | - | - | - |
| Regional/rural | -0.51 | 0.11 | 20.95 | 1 | <0.001 | 0.60 | 0.48 | 0.75 |
| **Gender** | | | | | | | | |
| Male (reference) | - | - | - | - | - | - | - | - |
| Female | -0.06 | 0.13 | 0.23 | 1 | 0.63 | 0.94 | 0.73 | 1.21 |
| **Time since diagnosis** | | | | | | | | |
| <6 months (reference) | - | - | - | - | - | - | - | - |
| 6–24 months | 0.45 | 0.18 | 8.46 | 1 | 0.004 | 1.57 | 1.16 | 2.13 |
| 2–5 years | 0.63 | 0.16 | 14.84 | 1 | <0.001 | 1.87 | 1.36 | 2.58 |
| >5 years | 0.76 | 0.19 | 15.10 | 1 | <0.001 | 2.13 | 1.45 | 3.11 |
| **Cancer stage** | | | | | | | | |
| Localized (reference) | - | - | - | - | - | - | - | - |
| Metastatic | -0.15 | 0.12 | 1.59 | 1 | 0.21 | 0.87 | 0.69 | 1.08 |
| Constant | -0.84 | 0.22 | 14.08 | 1 | <0.001 | 0.43 | | |

**Notes:** Variables entered into the model at each step: Step 1, time since study commencement, genitourinary cancer type; Step 2, age, location, gender; Step 3, time since diagnosis, cancer staging. Variable categories excluded from analysis: Non-binary/prefer not to say (gender); don't know/not applicable/other (cancer stage). Abbreviations: B, unstandardized coefficient; SE, standard error; df, degrees of freedom; OR, odds ratio; 95% CI, 95% confidence interval; AUD, Australian Dollars; K, 1000.

**Table S5.** Hierarchical multivariable logistic regression analysis of vaccinated status with head and neck cancer, compared with all other cancer types (n = 2545).

| | B | SE | Wald | df | p-value | OR | OR 95% CI | |
|---|---|---|---|---|---|---|---|---|
| | | | | | | | **Lower** | **Upper** |
| **Time since study commencement** | 0.03 | 0.002 | 213.06 | 1 | <0.001 | 1.03 | 1.03 | 1.04 |
| **Head and neck cancer type** | -0.57 | 0.24 | 5.52 | 1 | 0.02 | 0.56 | 0.35 | 0.91 |
| **Age (years)** | | | | | | | | |
| 18–49 (reference) | - | - | - | - | - | - | - | - |
| 50–69 | 0.44 | 0.14 | 10.44 | 1 | 0.001 | 1.56 | 1.19 | 2.03 |
| ≥70 | 1.27 | 0.17 | 55.96 | 1 | <0.001 | 3.57 | 2.56 | 4.98 |
| **Location** | | | | | | | | |
| Metropolitan (reference) | - | - | - | - | - | - | - | - |
| Regional/rural | -0.49 | 0.11 | 20.56 | 1 | <0.001 | 0.62 | 0.50 | 0.76 |
| **Gender** | | | | | | | | |
| Male (reference) | - | - | - | - | - | - | - | - |
| Female | -0.10 | 0.11 | 0.86 | 1 | 0.35 | 0.90 | 0.73 | 1.12 |
| **Current anti-cancer treatment** | | | | | | | | |
| Yes (reference) | - | - | - | - | - | - | - | - |
| No | 0.18 | 0.11 | 2.92 | 1 | 0.09 | 1.20 | 0.97 | 1.48 |
| Constant | -0.52 | 0.18 | 8.51 | 1 | 0.004 | 0.60 | | |

**Notes:** Variables entered into the model at each step: 1, time since study commencement, head and neck cancer; Step 2, age, location, gender; Step 3, current anti-cancer treatment. Variable categories excluded from analysis: Non-binary/prefer not to say (gender). Abbreviations: B, unstandardized coefficient; SE, standard error; df, degrees of freedom; OR, odds ratio; 95% CI, 95% confidence interval; AUD, Australian Dollars; K, 1000.

**Table S6.** Linear regression analysis predicting the Oxford COVID-19 Vaccine Hesitancy Scale summary score with sociodemographic and clinical characteristics.

| | | Step 1 | | | Step 2 | | |
|---|---|---|---|---|---|---|---|
| **Category (reference)** | **n** | **Adj. $R^2$** | **Adj. $R^2$** | **Δ Adj. $R^2$** | **B (SE)** | **t** | **p-value** |
| **Gender (Male)** | 2497 | 0.023 | 0.025 | 0.002 | | | |
| Female | | | | | 0.42 (0.20) | 2.15 | 0.032 |
| **Age (18–49 years)** | 2505 | 0.024 | 0.041 | 0.017 | | | |
| 50–69 | | | | | -1.35 (0.28) | -4.76 | <0.001 |
| ≥70 | | | | | -2.08 (0.30) | -6.87 | <0.001 |
| **Highest level of education (No formal /primary school/secondary school)** | 2504 | 0.024 | 0.027 | 0.003 | | | |
| Vocational/Trade | | | | | 0.40 (0.25) | 1.60 | 0.11 |
| University | | | | | -0.42 (0.22) | -1.86 | 0.063 |
| **Annual household income (AUD<50K)** | 2508 | 0.024 | 0.027 | 0.003 | | | |
| 50–100K | | | | | -0.42 (0.26) | -1.64 | 0.10 |
| 100K–150K | | | | | -0.16 (0.31) | -0.52 | 0.60 |
| >150K | | | | | -0.65 (0.33) | -1.96 | 0.051 |
| Prefer not to say | | | | | 0.45 (0.29) | 1.55 | 0.12 |
| **English as first language (Yes)** | 2507 | 0.024 | 0.029 | 0.005 | | | |
| No | | | | | 1.22 (0.33) | 3.69 | <0.001 |
| **Location (Metropolitan)** | 2508 | 0.024 | 0.025 | 0.001 | | | |
| Regional/rural | | | | | 0.48 (0.22) | 2.12 | 0.034 |
| **Cancer Type (All other cancer types – pooled)** | 2508 | | | | | | |
| Breast | | 0.024 | 0.024 | 0.00 | 0.32 (0.20) | 1.58 | 0.12 |
| Genitourinary | | 0.024 | 0.025 | 0.001 | -0.58 (0.25) | -2.33 | 0.02 |
| Gastrointestinal | | 0.024 | 0.024 | 0.00 | -0.19 (0.25) | -0.74 | 0.46 |
| Lung | | 0.024 | 0.024 | 0.00 | -0.45 (0.34) | -1.33 | 0.18 |
| Skin | | 0.024 | 0.024 | 0.00 | 0.70 (0.42) | 1.67 | 0.095 |
| Gynecological | | 0.024 | 0.023 | -0.001 | 0.26 (0.46) | 0.56 | 0.58 |
| Head and Neck | | 0.024 | 0.025 | 0.001 | 0.98 (0.51) | 1.91 | 0.056 |
| Other | | 0.024 | 0.023 | -0.001 | -0.14 (0.57) | -0.25 | 0.81 |
| **Cancer Stage (Localized)** | 2404 | 0.024 | 0.024 | 0.00 | | | |
| Metastatic | | | | | -0.13 (0.20) | -0.64 | 0.52 |
| **Time since diagnosis (<6 months)** | 2508 | 0.024 | 0.024 | 0.00 | | | |
| 6–24 months | | | | | -0.23 (0.31) | -0.74 | 0.46 |
| 2–5 years | | | | | -0.22 (0.31) | -0.71 | 0.48 |
| >5 years | | | | | -0.64 (0.34) | -1.86 | 0.062 |
| **Current anti-cancer treatment (Yes)** | 2508 | 0.024 | 0.023 | -0.001 | | | |
| No | | | | | -0.029 (0.20) | -0.15 | 0.88 |

**Notes:** Step 1, time since study commencement is the only predictor variable entered into the model; Step 2, the clinic-demographic is the predictor variable entered into the model. These variables were excluded due to <50 responses; Aboriginal and/or Torres Strait Islander status: non-binary/other gender, "other"' educational level. Cancer stage (don't know/other) were excluded for comparisons between localized and metastatic. Analyses were not undertaken for Aboriginal and/or Torres Strait Islander status, due to limited observations. Abbreviations: Adj. $R^2$, Adjusted $R^2$; B(SE), unstandardized coefficient (standard error); t, t-statistic; AUD, Australian Dollars; K, 1000.

**Table S7.** Hierarchical multivariable linear regression analysis of the Oxford COVID-19 Vaccine Hesitancy Scale score (n = 2500).

| **Step and predictor variable** | **B** | **SE** | **t** | **p-value** | **sr** | **Adj. $R^2$** | **ΔAdj. $R^2$** |
|---|---|---|---|---|---|---|---|
| Step 1 | | | | | | 0.024 | 0.024 |
| Constant | 11.30 | 0.22 | 52.46 | <0.001 | | | |
| **Time since study commencement** | -0.03 | 0.004 | -7.87 | <0.001 | -0.16 | | |
| Step 2 | | | | | | 0.052 | 0.028 |
| Constant | 12.72 | 0.35 | 36.76 | <0.001 | | | |
| **Time since study commencement** | -0.025 | 0.004 | -6.83 | <0.001 | -0.14 | | |
| **Age (years)** | | | | | | | |
| 18–49 (reference) | - | - | - | - | - | | |
| 50–69 | -1.43 | 0.28 | -5.03 | <0.001 | -0.1 | | |
| ≥70 | -2.22 | 0.31 | -7.24 | <0.001 | -0.14 | | |
| **Highest level of education** | | | | | | | |
| No formal/primary school/secondary school (reference) | - | - | - | - | - | | |
| Vocational/trade qualification | 0.17 | 0.25 | 0.69 | 0.49 | 0.014 | | |
| University | -0.81 | 0.23 | -3.61 | <0.001 | -0.07 | | |
| **English as first language** | | | | | | | |
| Yes (reference) | - | - | - | - | - | | |
| No | 1.24 | 0.33 | 3.78 | <0.001 | 0.08 | | |

**Notes:** Variables entered into the model at each step: Step 1, time since study commencement; Step 2, age, highest level of education, English as first language. Variable categories excluded from analysis: Other (Highest level of education). Abbreviations: B, unstandardized coefficient; SE, standard error; t, t-statistic; sr, semipartial correlation coefficient; Adj. $R^2$, Adjusted $R^2$.

**Table S8.** Hierarchical multivariable linear regression analysis of the Oxford COVID-19 Vaccine Hesitancy Scale score with genitourinary cancer type, compared with all cancer types (n = 2385).

| **Step and predictor variable** | **B** | **SE** | **t** | **p-value** | **sr** | **Adj. $R^2$** | **Δ Adj. $R^2$** |
|---|---|---|---|---|---|---|---|
| Step 1 | | | | | | 0.026 | 0.026 |
| Constant | 11.34 | 0.22 | 50.66 | <0.001 | | | |
| **Time since study commencement** | -0.03 | 0.004 | -7.67 | <0.001 | -0.16 | | |
| **Genitourinary cancer type** | -0.59 | 0.25 | -2.35 | 0.02 | -0.05 | | |
| Step 2 | | | | | | 0.05 | 0.024 |
| Constant | 12.60 | 0.40 | 31.56 | <0.001 | | | |
| **Time since study commencement** | -0.03 | 0.004 | -6.83 | <0.001 | -0.14 | | |
| **Genitourinary cancer type** | -0.14 | 0.30 | -0.47 | 0.64 | -0.01 | | |
| **Age (years)** | | | | | | | |
| 18–49 (reference) | - | - | - | - | - | | |
| 50–69 | -1.42 | 0.29 | -4.92 | <0.001 | -0.1 | | |
| ≥70 | -2.11 | 0.32 | -6.54 | <0.001 | -0.13 | | |
| **Gender** | | | | | | | |
| Male (reference) | - | - | - | - | - | | |
| Female | 0.09 | 0.24 | 0.39 | 0.70 | 0.008 | | |
| **Highest level of education** | | | | | | | |
| No formal/primary school/secondary school (reference) | - | - | - | - | - | | |
| Vocational/trade qualification | 0.25 | 0.26 | 0.96 | 0.34 | 0.02 | | |
| University | -0.74 | 0.23 | -3.20 | 0.001 | -0.07 | | |
| **English as first language** | | | | | | | |
| Yes (reference) | - | - | - | - | - | | |
| No | 1.00 | 0.34 | 2.91 | 0.004 | 0.06 | | |
| Step 3 | | | | | | 0.048 | -0.002 |
| Constant | 12.79 | 0.46 | 27.78 | <0.001 | | | |
| **Time since study commencement** | -0.03 | 0.004 | -6.72 | <0.001 | -0.14 | | |
| **Genitourinary cancer type** | -0.11 | 0.30 | -0.35 | 0.73 | -0.007 | | |
| **Age (years)** | | | | | | | |
| 18–49 (reference) | - | - | - | - | - | | |
| 50–69 | -1.40 | 0.29 | -4.84 | <0.001 | -0.10 | | |
| ≥70 | -2.09 | 0.32 | -6.45 | <0.001 | -0.13 | | |
| **Gender** | | | | | | | |
| Male (reference) | - | - | - | - | - | | |
| Female | 0.10 | 0.24 | 0.43 | 0.67 | 0.01 | | |
| **Highest level of education** | | | | | | | |
| No formal/primary school/secondary school (reference) | - | - | - | - | - | | |
| Vocational/trade qualification | 0.25 | 0.26 | 0.97 | 0.33 | 0.02 | | |
| University | -0.73 | 0.23 | -3.16 | 0.002 | -0.07 | | |
| **English as first language** | | | | | | | |
| Yes (reference) | - | - | - | - | - | | |
| No | 1.00 | 0.35 | 2.90 | 0.004 | 0.06 | | |
| **Time since diagnosis** | | | | | | | |
| <6 months (reference) | - | - | - | - | - | | |

| | | | | | |
|---|---|---|---|---|---|
| 6–24 months | -0.29 | 0.31 | -0.94 | 0.35 | -0.02 |
| 2–5 years | -0.24 | 0.32 | -0.76 | 0.45 | -0.02 |
| >5 years | -0.33 | 0.35 | -0.92 | 0.36 | -0.01 |
| **Cancer stage** | | | | | |
| Localized (reference) | - | - | - | - | - |
| Metastatic | 0.02 | 0.21 | 0.08 | 0.94 | 0.002 |

**Notes:** Variables entered into the model at each step: Step 1, time since study commencement; genitourinary cancer type; Step 2, age, gender, highest level of education, English as first language; Step 3, time since diagnosis, cancer stage. Variable categories excluded from analysis: Non-binary/prefer not to say (gender); other (highest educational level); don't know/not applicable/other (cancer stage). Abbreviations: B, unstandardized coefficient; SE, standard error; t, t-statistic; sr, semipartial correlation coefficient; Adj. $R^2$, Adjusted $R^2$.

**Table S9.** Linear regression predicting the Disease Influenced Vaccine Acceptance Scale-Six (DIVAS-6) Disease Complacency subscale score with sociodemographic and clinical characteristics.

| | | Step 1 | | | Step 2 | | |
|---|---|---|---|---|---|---|---|
| **Category (reference)** | **n** | **Adj. $R^2$** | **Adj. $R^2$** | **Δ Adj. $R^2$** | **B (SE)** | **t** | **p-value** |
| **Gender (Male)** | 2524 | 0.009 | 0.012 | 0.003 | | | |
| Female | | | | | -0.34 (0.12) | -2.76 | 0.006 |
| **Age (18–49 years)** | 2532 | 0.009 | 0.01 | 0.001 | | | |
| 50–69 | | | | | 0.21 (0.18) | 1.20 | 0.23 |
| ≥70 | | | | | 0.40 (0.19) | 2.08 | 0.04 |
| **Highest level of education (No formal /primary school/secondary school)** | 2529 | 0.009 | 0.008 | -0.001 | | | |
| Vocational/Trade | | | | | -0.04 (0.16) | -0.28 | 0.78 |
| University | | | | | -0.11 (0.14) | -0.78 | 0.44 |
| **Annual household income (AUD<50K)** | 2535 | 0.009 | 0.009 | 0.00 | | | |
| 50K–100K | | | | | -0.09 (0.16) | -0.57 | 0.57 |
| 100K–150K | | | | | -0.22 (0.19) | -1.13 | 0.26 |
| >150K | | | | | 0.02 (0.21) | 0.11 | 0.91 |
| Prefer not to say | | | | | 0.18 (0.18) | 1.01 | 0.31 |
| **English as first language (Yes)** | 2534 | 0.009 | 0.009 | 0.00 | | | |
| No | | | | | -0.05 (0.21) | -0.25 | 0.81 |
| **Location (Metropolitan)** | 2535 | 0.009 | 0.009 | 0.00 | | | |
| Regional/rural | | | | | -0.01 (0.14) | -0.05 | 0.96 |
| **Cancer Type (All other cancer types – pooled)** | 2535 | | | | | | |
| Breast | | 0.009 | 0.01 | 0.001 | -0.18 (0.13) | -1.45 | 0.15 |
| Genitourinary | | 0.009 | 0.015 | 0.006 | 0.60 (0.15) | 3.88 | <0.001 |
| Gastrointestinal | | 0.009 | 0.01 | 0.001 | -0.30 (0.16) | -1.92 | 0.06 |
| Lung | | 0.009 | 0.014 | 0.005 | -0.74 (0.21) | -3.54 | <0.001 |
| Skin | | 0.009 | 0.01 | 0.001 | 0.46 (0.27) | 1.72 | 0.09 |
| Gynecological | | 0.009 | 0.009 | 0.00 | -0.15 (0.28) | -0.55 | 0.58 |
| Head and Neck | | 0.009 | 0.01 | 0.001 | 0.53 (0.32) | 1.67 | 0.10 |
| Other | | 0.009 | 0.011 | 0.002 | 0.75 (0.35) | 2.11 | 0.04 |
| **Cancer Stage (Localized)** | 2426 | 0.009 | 0.017 | 0.008 | | | |
| Metastatic | | | | | -0.59 (0.13) | -4.66 | <0.001 |
| **Time since diagnosis (<6 months)** | 2535 | 0.009 | 0.008 | -0.001 | | | |
| 6–24 months | | | | | -0.05 (0.19) | -0.25 | 0.80 |
| 2–5 years | | | | | -0.05 (0.20) | -0.24 | 0.81 |
| >5 years | | | | | 0.07 (0.21) | 0.34 | 0.73 |
| **Current anti-cancer treatment (Yes)** | 2535 | 0.009 | 0.016 | 0.007 | | | |
| No | | | | | 0.51 (0.12) | 4.19 | <0.001 |

**Notes:** Step 1 = time since study commencement is the only predictor variable entered into the model; Step 2 = the clinic-demographic is the predictor variable entered into the model. These variables were excluded due to <50 responses; Aboriginal and/or Torres Strait Islander status: non-binary/other gender, "other"' educational level. Cancer stage (don't know/other) were excluded for comparisons between localized and metastatic. Abbreviations: Adj. $R^2$, Adjusted $R^2$; B(SE), unstandardized coefficient (standard error); t, t-statistic; AUD, Australian Dollars; K, 1000.

**Table S10.** Linear regression predicting the Disease Influenced Vaccine Acceptance Scale-Six (DIVAS-6) Vaccine Vulnerability subscale score with sociodemographic and clinical characteristics.

| | | Step 1 | | | Step 2 | | |
|---|---|---|---|---|---|---|---|
| **Category (reference)** | **n** | **Adj. $R^2$** | **Adj. $R^2$** | **Δ Adj. $R^2$** | **B (SE)** | **t** | **p-value** |
| **Gender (Male)** | 2316 | 0.005 | 0.015 | 0.01 | | | |
| Female | | | | | 0.74 (0.15) | 5.02 | <0.001 |
| **Age (18–49 years)** | 2325 | 0.005 | 0.03 | 0.025 | | | |
| 50–69 | | | | | -0.69 (0.21) | -3.31 | 0.001 |
| ≥70 | | | | | -1.65 (0.23) | -7.28 | <0.001 |
| **Highest level of education (No formal /primary school/secondary school)** | 2324 | 0.005 | 0.015 | 0.01 | | | |
| Vocational/Trade | | | | | 0.05 (0.19) | 0.28 | 0.78 |
| University | | | | | -0.73 (0.17) | -4.32 | <0.001 |
| **Annual household income (AUD<50K)** | 2328 | 0.005 | 0.017 | 0.012 | | | |
| 50-100K | | | | | -0.38 (0.20) | -1.96 | 0.051 |
| 100K-150K | | | | | 0.07 (0.23) | 0.32 | 0.75 |
| >150K | | | | | -0.74 (0.25) | -3.00 | 0.003 |
| Prefer not to say | | | | | 0.67 (0.22) | 3.03 | 0.002 |
| **English as first language (Yes)** | 2327 | 0.005 | 0.017 | 0.012 | | | |
| No | | | | | 1.36 (0.25) | 5.38 | <0.001 |
| **Location (Metropolitan)** | 2328 | 0.005 | 0.005 | 0.00 | | | |
| Regional/rural | | | | | 0.14 (0.17) | 0.79 | 0.43 |
| **Cancer Type (All other cancer types – pooled)** | 2328 | | | | | | |
| Breast | | 0.005 | 0.005 | 0.00 | 0.15 (0.15) | 0.98 | 0.33 |
| Genitourinary | | 0.005 | 0.017 | 0.012 | -1.00 (0.19) | -5.36 | <0.001 |
| Gastrointestinal | | 0.005 | 0.007 | 0.002 | 0.41 (0.19) | 2.17 | 0.03 |
| Lung | | 0.005 | 0.007 | 0.002 | 0.58 (0.26) | 2.25 | 0.03 |
| Skin | | 0.005 | 0.005 | 0.00 | -0.27 (0.32) | -0.83 | 0.40 |
| Gynecological | | 0.005 | 0.007 | 0.002 | 0.87 (0.36) | 2.43 | 0.02 |
| Head and Neck | | 0.005 | 0.006 | 0.001 | -0.50 (0.40) | -1.26 | 0.21 |
| Other | | 0.005 | 0.005 | 0.00 | 0.16 (0.44) | 0.37 | 0.71 |
| **Cancer Stage (Localized)** | 2237 | 0.004 | 0.011 | 0.007 | | | |
| Metastatic | | | | | 0.62 (0.15) | 4.05 | <0.001 |
| **Time since cancer diagnosis (<6 months)** | | 0.005 | 0.016 | 0.011 | | | |
| 6–24 months | | | | | -0.43 (0.23) | -1.89 | 0.06 |
| 2–5 years | | | | | -0.75 (0.24) | -3.15 | 0.002 |
| >5 years | | | | | -1.25 (0.26) | -4.84 | <0.001 |
| **Current anti-cancer treatment (Yes)** | 2328 | 0.005 | 0.017 | 0.012 | | | |
| No | | | | | -0.79 (0.15) | -5.33 | <0.001 |

**Notes:** Step 1 = time since study commencement is the only predictor variable entered into the model; Step 2 = the clinic-demographic is the predictor variable entered into the model. These variables were excluded due to <50 responses; Aboriginal and/or Torres Strait Islander status: non-binary/other gender, "other"' educational level. Cancer stage (don't know/other) were excluded for comparisons between localized and metastatic. Abbreviations: Adj. $R^2$, Adjusted $R^2$; B(SE), unstandardized coefficient (standard error); t, t-statistic; AUD, Australian Dollars; K, 1000.

**Table S11.** Hierarchical multivariable linear regression of the DIVAS-6 Disease Complacency subscale score with genitourinary cancer type, compared with all other cancer types (n = 2412).

| Step and predictor variable | B | SE | t | p-value | sr | Adj. $R^2$ | Δ Adj. $R^2$ |
|---|---|---|---|---|---|---|---|
| Step 1 | | | | | | 0.015 | 0.015 |
| Constant | 6.84 | 0.14 | 48.63 | <0.001 | | | |
| **Time since study commencement** | -0.01 | 0.002 | -4.74 | <0.001 | -0.10 | | |
| **Genitourinary cancer type** | 0.63 | 0.16 | 3.97 | <0.001 | 0.08 | | |
| Step 2 | | | | | | 0.014 | -0.001 |
| Constant | 6.75 | 0.23 | 29.12 | <0.001 | | | |
| **Time since study commencement** | -0.01 | 0.002 | -4.80 | <0.001 | -0.10 | | |
| **Genitourinary cancer type** | 0.53 | 0.19 | 2.80 | 0.005 | 0.06 | | |
| **Age (years)** | | | | | | | |
| 18–49 (reference) | - | - | - | - | - | | |
| 50–69 | 0.16 | 0.18 | 0.88 | 0.38 | 0.02 | | |
| ≥70 | 0.25 | 0.20 | 1.22 | 0.22 | 0.03 | | |
| **Gender** | | | | | | | |
| Male (reference) | - | - | - | - | - | | |
| Female | -0.09 | 0.15 | -0.58 | 0.57 | -0.01 | | |
| Step 3 | | | | | | 0.026 | 0.012 |
| Constant | 7.09 | 0.28 | 25.73 | <0.001 | | | |
| **Time since study commencement** | -0.01 | 0.002 | -5.17 | <0.001 | -0.11 | | |
| **Genitourinary cancer type** | 0.61 | 0.19 | 3.15 | 0.002 | 0.06 | | |
| **Age (years)** | | | | | | | |
| 18–49 (reference) | - | - | - | - | - | | |
| 50–69 | 0.20 | 0.18 | 1.10 | 0.27 | 0.02 | | |
| ≥70 | 0.28 | 0.20 | 1.38 | 0.17 | 0.03 | | |
| **Gender** | | | | | | | |
| Male (reference) | - | - | - | - | - | | |
| Female | -0.19 | 0.15 | -1.22 | 0.22 | -0.03 | | |
| **Time since diagnosis** | | | | | | | |
| <6 months (reference) | - | - | - | - | - | | |
| 6–24 months | -0.02 | 0.20 | -0.1 | 0.92 | -0.002 | | |
| 2–5 years | 0.005 | 0.20 | 0.02 | 0.98 | 0 | | |
| >5 years | 0.06 | 0.23 | 0.28 | 0.78 | 0.006 | | |
| **Cancer stage** | | | | | | | |
| Localized (reference) | - | - | - | - | - | | |
| Metastatic | -0.74 | 0.13 | -5.66 | <0.001 | -0.12 | | |

***Notes:*** Variables entered into the model at each step: Step 1, time since study commencement, genitourinary cancer type; Step 2, age, gender; Step 3, time since diagnosis, cancer stage. Variable categories excluded from analysis: Non-binary/prefer not to say (gender); don't know/not applicable/other (cancer stage). Abbreviations: B, unstandardized coefficient; SE, standard error; t, t-statistic; sr, semipartial correlation coefficient; Adj. $R^2$, Adjusted $R^2$.

**Table S12.** Hierarchical multivariable linear regression analysis of the DIVAS-6 Vaccine Vulnerability subscale score with genitourinary cancer type, compared with all other cancer types (n = 2221).

| | B | SE | t | p-value | sr | Adj. $R^2$ | Δ Adj. $R^2$ |
|---|---|---|---|---|---|---|---|
| Step 1 | | | | | | 0.017 | 0.017 |
| Constant | 8.91 | 0.17 | 53.06 | <0.001 | | | |
| **Time since study commencement** | -0.01 | 0.003 | -3.02 | 0.003 | -0.06 | | |
| **Genitourinary cancer type** | -1.04 | 0.19 | -5.47 | <0.001 | -0.12 | | |
| Step 2 | | | | | | 0.043 | 0.026 |
| Constant | 9.14 | 0.28 | 33.24 | <0.001 | | | |
| **Time since study commencement** | -0.006 | 0.003 | -2.17 | 0.03 | -0.05 | | |
| **Genitourinary cancer type** | -0.55 | 0.22 | -2.46 | 0.014 | -0.05 | | |
| **Age (years)** | | | | | | | |
| 18–49 (reference) | - | - | - | - | - | | |
| 50–69 | -0.55 | 0.21 | -2.58 | 0.01 | -0.06 | | |
| ≥70 | -1.35 | 0.24 | -5.66 | <0.001 | -0.12 | | |
| **Gender** | | | | | | | |
| Male (reference) | - | - | - | - | - | | |
| Female | 0.26 | 0.18 | 1.43 | 0.15 | 0.03 | | |
| **English as first language** | | | | | | | |
| Yes (reference) | - | - | - | - | - | | |
| No | 1.16 | 0.26 | 4.42 | <0.001 | 0.09 | | |
| Step 3 | | | | | | 0.068 | 0.025 |
| Constant | 9.40 | 0.33 | 28.52 | <0.001 | | | |
| **Time since study commencement** | -0.004 | 0.003 | -1.55 | 0.12 | -0.03 | | |
| **Genitourinary cancer type** | -0.55 | 0.23 | -2.43 | 0.02 | -0.05 | | |
| **Age (years)** | | | | | | | |
| 18–49 (reference) | - | - | - | - | - | | |
| 50–69 | -0.55 | 0.21 | -2.64 | 0.008 | -0.06 | | |
| ≥70 | -1.34 | 0.24 | -5.69 | <0.001 | -0.12 | | |
| **Gender** | | | | | | | |
| Male (reference) | | | | | | | |
| Female | 0.33 | 0.18 | 1.84 | 0.07 | 0.04 | | |
| **English as first language** | | | | | | | |
| Yes (reference) | - | - | - | - | - | | |
| No | 1.07 | 0.26 | 4.13 | <0.001 | 0.09 | | |
| **Time since diagnosis** | | | | | | | |
| <6 months (reference) | - | - | - | - | - | | |
| 6–24 months | -0.28 | 0.23 | -1.20 | 0.23 | -0.03 | | |
| 2–5 years | -0.57 | 0.24 | -2.37 | 0.02 | -0.05 | | |
| >5 years | -0.96 | 0.27 | -3.61 | <0.001 | -0.08 | | |
| **Cancer stage** | | | | | | | |
| Localized (reference) | - | - | - | - | - | | |
| Metastatic | 0.80 | 0.16 | 4.95 | <0.001 | 0.11 | | |
| **Current anti-cancer treatment** | | | | | | | |
| Yes (reference) | - | - | - | - | - | | |
| No | -0.56 | 0.16 | -3.56 | <0.001 | -0.08 | | |

**Notes:** Variables entered into the model at each step: Step 1, time since study commencement, genitourinary cancer type; Step 2, age, gender, English as first language; Step 3, time since diagnosis, cancer stage, current anti-cancer treatment. Variable categories excluded from analysis: Non-binary/prefer not to say (gender); don't know/not applicable/other (cancer stage). Abbreviations: B, unstandardized coefficient; SE, standard error; t, t-statistic; sr, semipartial correlation coefficient; Adj. $R^2$, Adjusted $R^2$.
